# Artificial Intelligence-Assisted Loop Mediated Isothermal Amplification (ai-LAMP) for Rapid Detection of SARS-CoV-2

**DOI:** 10.1101/2020.07.08.20148999

**Authors:** Mohammed A Rohaim, Emily Clayton, Irem Sahin, Julianne Vilela, Manar E Khalifa, Mohammad Q Al-Natour, Mahmoud Bayoumi, Aurore Poirier, Manoharanehru Branavan, Mukunthan Tharmakulasingam, Nouman S Chaudhry, Ravinder Sodi, Amy Brown, Peter Burkhart, Wendy Hacking, Judy Botham, Joe Boyce, Hayley Wilkinson, Craig Williams, Michelle Bates, Roberto La Ragione, Wamadeva Balachandran, Anil Fernando, Muhammad Munir

## Abstract

Until vaccines and effective therapeutics become available, the practical solution to transit safely out of the current coronavirus disease 19 (CoVID-19) lockdown may include the implementation of an effective testing, tracing and tracking system. However, this requires a reliable and clinically validated diagnostic platform for the sensitive and specific identification of SARS-CoV-2. Here, we report on the development of a *de novo*, high-resolution and comparative genomics guided reverse-transcribed loop-mediated isothermal amplification (LAMP) assay. To further enhance the assay performance and to remove any subjectivity associated with operator interpretation of results, we engineered a novel hand-held smart diagnostic device. The robust diagnostic device was further furnished with automated image acquisition and processing algorithms, and the collated data was processed through artificial intelligence (AI) pipelines to further reduce the assay run time and the subjectivity of the colorimetric LAMP detection. This advanced AI algorithm-implemented LAMP (ai-LAMP) assay, targeting the RNA-dependent RNA polymerase gene, showed high analytical sensitivity and specificity for SARS-CoV-2. A total of ∼200 coronavirus disease (CoVID-19)-suspected NHS patient samples were tested using the platform and it was shown to be reliable, highly specific and significantly more sensitive than the current gold standard qRT-PCR. Therefore, system could provide an efficient and cost-effective platform to detect SARS-CoV-2 in resource-limited laboratories.

## 1. Introduction

A cluster of new pneumonia cases was reported to the World Health Organization (WHO) in late 2019 from Wuhan, Hubei Province of China. The causative agent was named as severe acute respiratory syndrome coronavirus 2 (SARS-CoV-2) and led to a global pandemic [1–3]. While the major impact of SARS-CoV-2 was attributed to frail and elderly people with co-morbidities, coronavirus disease 2019 (CoVID-19) was mainly spread by asymptomatic or mildly symptomatic patients [2]. Due to their high mutation rates and recombination events, coronaviruses can infect a range of animal species including humans, avian, rodents, carnivores, chiropters and other mammals [4]. Prior to the emergence of SARS-CoV-2, a total of six different coronaviruses had been reported to infect humans, including HCoV-229E, HCoV-OC43, HCoV-NL63, HCoV-HKU1, MERS and SARS-CoV-1 (also known as classical SARS). The SARS-CoV-2 belongs to the β-coronavirus of the group 2B within the family of *Coronaviridae* [3].

The SARS-CoV-2 shares a high level of genetic similarity (up to 96%) with coronaviruses originating from bats [3]. The genome of β-coronavirus encodes for the replicase complex (ORF1ab), spike (S), envelope (E), membrane (M) and nucleoprotein (N) genes in addition to the several non-structural and accessory proteins in the order from 5’-untranslated to 3’-untranslated regions [3]. Owing to the nature of viral genetics, the N gene is the most transcribed and highly conserved gene within the *Coronaviridae* family and has been a major target for both antigen and antibodies diagnostics. Across the genome, the RNA-dependent RNA polymerase (RdRP), encoded by the ORF1b gene segment, presents a high level of intra-group conservation and therefore is an ideal target for a diagnostic application [5, 6].

As evident by previous coronaviruses associated pandemics and other viral diseases, a highly specific, sensitive and easily deployable diagnostic is critical for the identification, tracing, rationalizing control measures, documentation of symptomatic and asymptomatic carriers [7-12]. Additionally, due to the unavailability of the registered vaccines or effective therapeutics, rapid and reliable diagnostics are of paramount importance to curtail SARS-CoV-2 infection. Because of shortcomings associated with the virus isolation (time consuming and required containment) and cross-reactivities of antigen and antibodies assay, several real-time reverse transcription-polymerase chain reactions (qRT-PCR) and reverse-transcription loop mediated isothermal amplification (RT-LAMP) assays have been developed, validated and commercialized as useful laboratory diagnostics for the detection of SARS-CoV-2 [13]. However, the majority of these assays are time-consuming and require laboratory-intense instrumentation. Furthermore, they are unable to meet the current unprecedented rapid growth and demand for testing a large proportion of the population, identification of asymptomatic carriers and contact tracing.

Though qRT-PCR remains the gold standard for the diagnosis of SARS-CoV-2, RT-LAMP assays have been demonstrated to produce diagnostic results with increased sensitivity and specificity [14]. Furthermore, its ability to tolerate PCR inhibitors eliminates the need for laborious RNA extraction and purification methodologies [15, 16]. Several platforms capable of performing LAMP assays in the field have previously been documented [17]. However, most platforms have employed fluorescence detection with integrated optical units or a smart phone dock to achieve detection [18, 19]. Similarly, for colorimetric LAMP assays, smart phone cameras or user interpretation of the colour changes were used to achieve detection [20, 21]. The fully integrated real-time fluorescence-based platforms are expensive, and the smartphone-based platforms are only designed for specific smartphone models. Therefore, to fulfil the need for a standalone colorimetric isothermal nucleic acid amplification platform [22], we have developed an ultra-low-cost molecular diagnostic device with an integrated single-board computer, imaging camera, artificial intelligence-based image processing algorithm and mobile app.

In this study, we developed a high-resolution comparative genomics analysis-guided novel RT-LAMP assay for the specific and sensitive detection of SARS-CoV-2 in comparison to WHO recommended qRT-PCR assays. In order to provide a simple “sample-to-answer workflow”, an ultra-low-cost and user-friendly diagnostic platform was engineered and further enhanced with a module for automated image acquisition and processing. Artificial intelligence-guided assessment of the LAMP assay provided faster detection of colour changes in the LAMP reaction, further enhancing the assay performance and thus reducing the potential for human error in results interpretation. Finally, the assay was validated on RNA extracted from clinical samples from SARS-CoV-2 suspected patients to demonstrate the real-life applicability.

## 2. Experimental Section

### 2.1. Ethics statement

This study was conducted in accordance with the University Human ethics guidelines and received a favourable review from the Faculty of Health and Medicine Research Ethics Committee (FHMREC) of Lancaster University - reference number FHMREC19112. The study was exempt from requiring specific patient consent, as it only involved the use of extracted RNA and existing collections of data or records that contained non-identifiable data about human patients.

### 2.2. Cells and viruses

Vero cells and MDCK cells were cultured in Dulbecco’s modified Eagle’s medium (DMEM) (Gibco, Carlsbad, CA) supplemented with 10% inactivated foetal bovine serum (FBS) (Gibco), 2 mM l-glutamine (Gibco) and 100U/mL penicillin/streptomycin (Gibco) at 37°C in 5% CO2. Influenza A virus (A/chicken/Pakistan/UDL-01/2008(H9N2), Newcastle disease virus strain LaSota and Infectious bronchitis virus strain H120, Vesicular stomatitis virus (VSV) and Sendai virus (SeV) were propagated and used to determine the specificity of the LAMP. All viruses except influenza were titrated on Vero and MDCK cells, respectively by the standard plaque assay.

### 2.3. In silico nucleotide sequence comparisons and primer design

To design specific LAMP primer sets for the detection of SARS-CoV-2, all available complete genome sequences were downloaded from GISAID Initiative (https://www.gisaid.org/), aligned and the conserved part was selected and used as the template of LAMP primer design. To find out an efficient primer set, three sets of specific LAMP primers were hand-picked and validated using PrimerExplorer V5 software (http://primerexplorer.jp/elamp4.0.0/index.html). Primers were validated using BLAST software (http://www.ncbi.nlm.gov/BLAST) to ensure their specificity.

### 2.4. Cloning and in vitro transcription of RdRP target gene

The coding sequence of SARS-CoV-2 RdRp gene was chemically synthesized and cloned into *pVAX1* plasmid (Invitrogen, Carlsbad, USA) between *Kpn*I and *Not*I restriction sites. The plasmid was propagated in DH5α cells and purified using MiniPrep Qiagen Kits. The linearized plasmid with pVAX1-RdRP was used for *in vitro* transcription using T7 RiboMAX™ Express Large-Scale RNA Production System (Promega, USA). The copy number of *in vitro* transcribed RNA was calculated from RNA concentration measured with NanoDrop™ 2000c Spectrophotometers (Thermo, USA) in triplicate. RNA products were then purified using the RNeasy MinElute Cleanup Kit (Qiagen, Valencia, CA, USA). A standard curve was generated using dilutions of the standard *in vitro* transcribed RNAs using *SuperScript III Platinum One*-*Step qRT*-*PCR Kit* as per the manufacturer’s protocol (Invitrogen, Carlsbad, USA) using CFX384 Touch Real-Time PCR Detection System is (Applied Biosystems, USA).

### 2.5. Clinical sample processing and spiking with miR-cel-miR-39-3p RNA

A total of 199 nasopharyngeal swabs were individually collected from CoVID-19 suspected patients, through routine NHS collection procedure for COVID-19 screening. These samples were stored and transported in the virus transport media (VTM) to the NHS diagnostic laboratory at Lancaster University, UK. All samples were individually spiked with 50 pmol/L of synthesized *Caenorhabditis elegans* miR-cel-miR-39-3p (Applied Biosystems/Thermo-Fisher Scientific, UK). The miR-cel-miR-39-3p RNA lacked any sequence homology to human or viral gene and thus present an ideal RNA extraction control. Total RNA including miRNAs was extracted using 140 μL of the spiked-VTM by the commercial QIAampViral RNA Mini kit (Qiagen, Valencia, California). The miR-cel-miR-39-3p RNA was used to serve as an internal control to monitor extraction efficiency and used for data normalization. The final RNA yield and purity were determined by the A260/A280 ratio measured by a NanoDrop ND-1000 spectrophotometer (NanoDrop Technologies/Thermo-Fisher Scientific, UK) with a ratio of 1.80 to 2.00 indicative of good RNA purity. The isolated RNA was stored at −80 °C for further use.

### 2.6. Real-time fluorescent-based quantitative PCR

Suspected SARS-CoV-2 clinical samples were tested for positivity by qRT-PCR. Briefly, RNA was extracted from Viral Transport Media using the QIAamp Viral RNA Mini kit (Qiagen, Valencia, California) following the manufacturer instructions. The qRT-PCR was conducted using the *SuperScript III Platinum One*-*Step qRT*-*PCR Kit* as per the manufacturer’s protocol (Invitrogen Carlsbad, USA) in the CFX384 Touch Real-Time PCR Detection System (Biorad, USA), according to the cycling protocol. The reaction was performed using the specific primer set RdRpF; RdRpR and FAM-labelled probe or NP-F; NP-R and ROX labelled probes designed to detect SARS-CoV2. The 25-µl PCR reaction consists of 12.5 µl 2X Reaction Mix, 0.2 µM of each primer, and 0.1 µM probe, 0.5 µl of SuperScript® III RT/Platinum® Taq Mix, 5 µl of RNA sample and nuclear free water. The cycling program was performed in the CFX384 Touch Real-Time PCR Detection System is (Applied Biosystems, USA), according to the cycling protocol. The amount of viral RNA in each sample was estimated by comparing the cycle threshold values (Ct) to the standard curve made by serial 10-fold serial dilutions of previously titrated *in vitro* transcribed RdRP gene.

### 2.7. ai-LAMP assay performance

All experiments for LAMP were run in triplicate. The LAMP reactions were performed using WarmStart™Colorimetric LAMP 2X Master Mix (New England Biolabs). A 10X primer mix (FIP, 16 µM; BIP, 16 µM; F3, 2 µM; B3, 2 µM; LF, 4 µM; LB, 4 µM) was prepared. A 25 µl reaction mixture (12.5 µl 2X MasterMix; 2.5 µl 10X primer mix; 2.5 µl RNA and 7.5 µl DNase & RNase-free molecular grade water) was mixed homogeneously and centrifuged. The LAMP assays were performed in a thermocycler (MJResearch) at 65°C for 30 min or in the engineered device (**Figure 4A**). Colour change was observed directly by the naked eye or through AI image processing, and agarose gel electrophoresis was performed to confirm the results. The completion of amplification was indicated by the colour in the tube, wherein yellow was considered positive and pink was regarded as negative. All amplicons were confirmed by 2% agarose gel electrophoresis.

### 2.8. Artificial intelligence based test-tube colour detection

A loop-mediated isothermal amplification (LAMP) assay based COVID-19 test device was developed to capture the COVID-19 test results in 30 minutes, based on colour changes. Artificial intelligence (AI) based colour detection was proposed to identify colour changes considering different lighting issues and to reduce the test running time less than 30 minutes. Images were acquired from the COVID-19 test kit which carried 8 test-tubes including NTC (negative test control) and PTC (positive test control) for every 20 seconds during the test operation. Each image was cropped into separate tubes using template matching approach and labelled manually based on their colour.

### 2.9. Analytical specificity and analytical sensitivity of the LAMP assay

The designed RdRp primer sets for LAMP to detect SARS-CoV-2 were validated for their specificity by testing the cross-reactivity with other viruses, including influenza A virus, Vesicular stomatitis virus (VSV), Sendai virus (SeD), infectious bronchitis virus (IBV) and Newcastle disease virus (NDV). Likewise, the developed LAMP assay was evaluated to test the primers set sensitivity in a serially diluted standard RNA template prepared by tenfold serial dilutions. The amplification patterns were observed for each dilution to determine the lowest amount of absolute RNA template required for detectable amplification. The degree of colour intensity of the amplified product as well as the observed electrophoretic pattern during gel electrophoresis was used for the analysis of LAMP amplification.

### 2.10. Quantitative real time PCR for miR-cel-miR-39-3p RNA

In order to determine the RNA extraction efficiency, the extracted RNA was reverse transcribed using a commercially available kit (Applied Biosystems/Thermo-Fisher Scientific, UK) using miR-specific stem-loop primers as per manufacturer instructions. A total of 5 μL of the sample was added to a 96-well plate together with 10 μL reaction mixture (MasterMixTM) containing along with MultiscribeTM reverse transcriptase (50 U/μL), and 0.19 μL RNAase inhibitor (20 U/μL). The RT reaction was performed at 16 °C for 30 min, followed by 42 °C for 30 min, and 85 °C for 5 min and was finally kept at 4 °C. A NTC was considered in every individually run reaction to identify any unspecific amplification. The RT products were quantified immediately by qPCR using TaqManTM MicroRNA assays (Applied Biosystems/Thermo-Fisher Scientific, UK) in a 96 well plate using the 7900HT Fast Real-Time PCR System (Applied Biosystems, UK) as we described before [23]. The quantification cycle (Cq) was determined with instrument default threshold settings (10 SDs above the mean fluorescence of the baseline cycle).

### 2.11. Statistical analysis

A total of 200 sample size was calculated to assess the performance of the LAMP assay. GraphPad Prism Software version 6.01 for Mac (GraphPad Software, La Jolla, California, USA) was used for graphs generation. The LAMP detection sensitivity and specificity were calculated using the chi-squared test. TPR (true positive rate), TNR (true negative rate), FPR (false positive rate), FNR (false negative rate) were calculated according to the following equations: TPR= TP/(TP+FN). TNR=TN/(FP+TN). FNR=FN/(TP+FN). FPR=FP/(FP+TN). TP: total number of true positives. TN: total number of true negatives. TN: total number of true negatives. FN: total number of false negatives.

## 3. Results

### 3.1. High resolution conversation analysis of SARS-CoV-2 to guide oligo design

It is imperative to critically assess the evolving nature of viruses in identifying conserved gene signatures and guiding the selection of the most appropriate primers. In order to identify important genomic loci, we downloaded and aligned all the available full-length genomes with high coverage sequences (*n*=22858) of SARS-CoV-2 by Multiple Alignment using Fast Fourier Transform (MAFFT) [24]. We then compiled *in house* R-code (available on request) to determine the single nucleotide-based genetic conservation across the length of ∼30kb genome. The analysis of the aligned dataset of all genomes in the RStudio generated a total of 18GB high-resolution nucleotide-by-nucleotide score from 0.0 to 1.0 (1.0 being the highly conserved and 0.0 being the highly divergent). Plotting the assessed genetic divergence, at a cut-off point of 90% similarity along with the genome of the SARS-CoV-2, identified sharp divergence at multiple loci (Figure 1A). However, most of the genomes maintained high conservation. The divergence at the 5’ and 3’ ends was primarily due to length heterogeneity, which may be partly as a result of sequencing artifact or potentially coronaviruses ragged termini (Figure 1B). Owing to high divergence, a stretch of sequence (∼400 nucleotides, numbering corresponds to the complete genome of strain SARS-CoV-2/human/USA/VA-DCLS-0285/2020 strain, GenBank Accession Number: MT558705.1) spanning the start of the ORF1b, which encode for viral RNA-dependent RNA polymerase (RdRP), was targeted to design oligos for the LAMP assay. Additionally, this specific target genomic locus was adjacent to oligos recommended by the World Health Organization (WHO) and Public Health England (PHE) for real-time RT-PCR-based routine identification of CoVID-19 patients, further allowing direct and comparable evaluation of real-time RT-PCR with *de novo* developed LAMP assay (Figure 1C).

**Figure 1.**
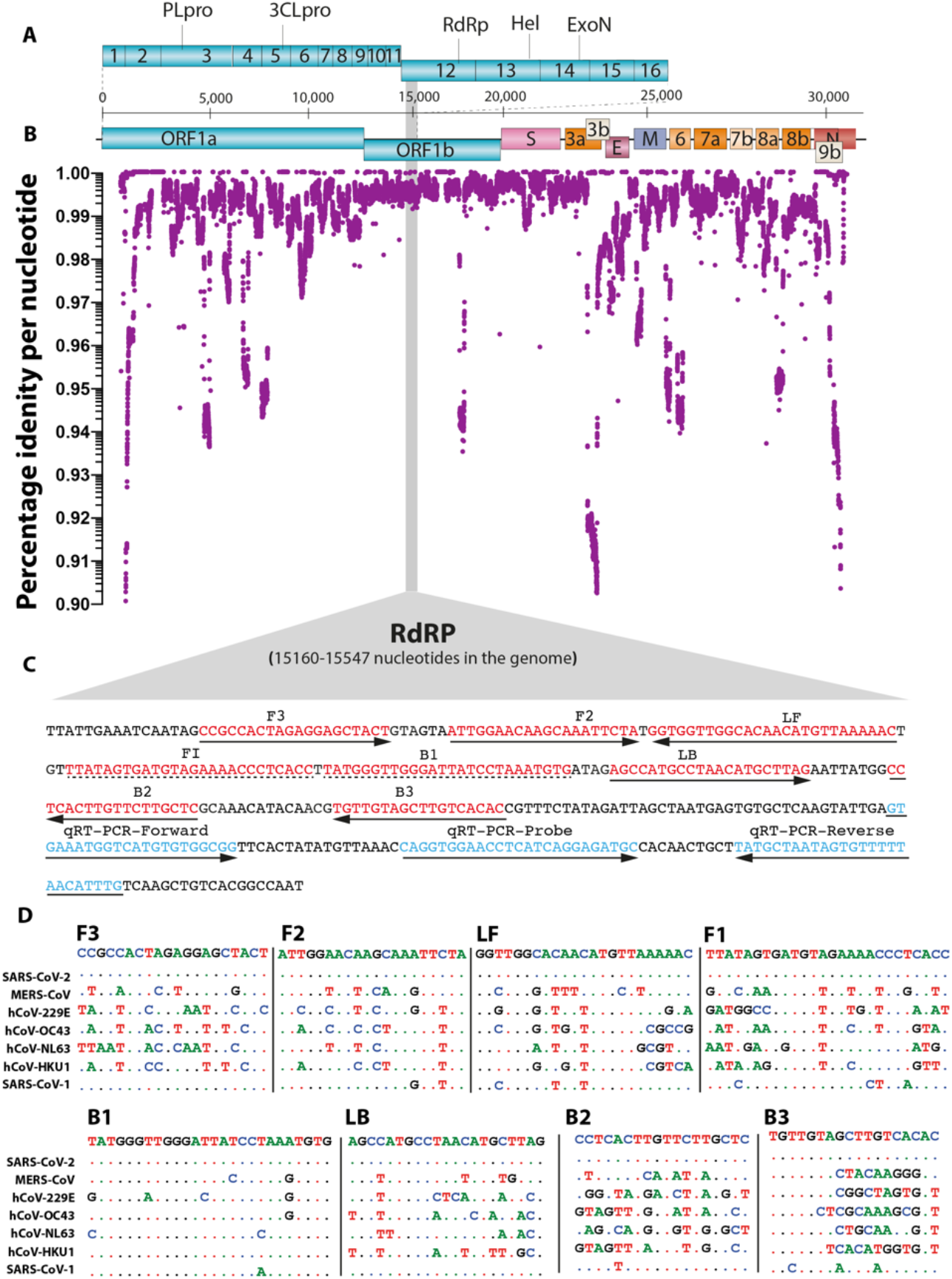
In silico analysis of SARS-CoV-2 and primer design. (**A**) Genome organization of SARS-CoV-2. Scale represents an approximate position of the genome whereas ORF1a and b are expanded to show internal gene organization. (**B**) Level of gene identity across the genome of the SARS-CoV-2. Identity less than 90% is not shown. (**C**) Primer location in the RdRP gene of SARS-COV-2 is shown. Red coloured sequences represent LAMP primers whereas blue coloured sequences are primers and probes used in the qRT-PCR. (**D**) Comparative sequence identity using the primers against different human coronaviruses compared to the reference SARS-CoV-2 sequence; dots represent identical nucleotides.

The conserved region of the RdRP gene with the lowest mutation frequencies was used as a template to manually design three sets of basic LAMP primers and selected with PrimerExplorer V5 for appropriate primer lengths, loop selection and melting temperature optimization (Figure 1C). In order to preclude the non-specific amplification of common coronaviruses, efforts were made to design primers in the regions where there is a high level of divergence among more than 3 of the 6 total primers in a specific set. Amongst the most suitable targets, the primers with high scores were aligned with MERS-CoV, hCoV-229E, hCoV-OC43, hCoV-NL63, hCoV-HKU1 and SARS-CoV-1 (Figure 1D). These selected primers were used for further validation and screening.

### 3.2. Determination of the limit of detection of the LAMP assay using biochemically synthesized RNA

In order to assess the robustness of the primers, we used a fully identical *in vitro* transcribed target RNA unanimously spanning the length of the RdRP-gene based LAMP and qRT-PCR target regions. The pre-determined copy numbers of the biochemically synthesised RNA were 10-fold serially diluted from 10^7^ copies to 0 copies of the target gene per reaction. To determine the analytical sensitivity of the assays, we first evaluated their limits of detection (LoD) for both qRT-PCR and LAMP assays. The LoD of the qRT-PCR was 10 copies as evident from the relative fluorescence units (Figure 2A) and electrophoreses of the amplified products (Figure 2B). The standard curve generated by the RdRP-based qRT-PCR was linear and generated a coefficient of correlation (R^2^) = 0.9481 and a slope of -2.6509 (Figure 2C). Melting curve analysis revealed the specificity of primers for the target gene sequence, as all the amplified products showed a uniform melting temperature (Tm) of ∼75.10°C and specific amplification patterns (Figure 2B and data not shown). Compared to the qRT-PCR assay, the LoD for the LAMP which targeted the same RdRP gene was 1 log unit higher (10^2^ copies/reaction) (Figure 2D, upper panel) as assessed by visual observation of the LAMP reaction, where positive reactions turned yellow and negative reactions remained pink when observed by the naked eye. To further confirm the specific amplification of the target region, the gradient LAMP products were visualized by DNA staining and gel electrophoresis for the amplified product, further confirming the detection limit of LAMP (Figure 2D, lower panel).

**Figure 2:**
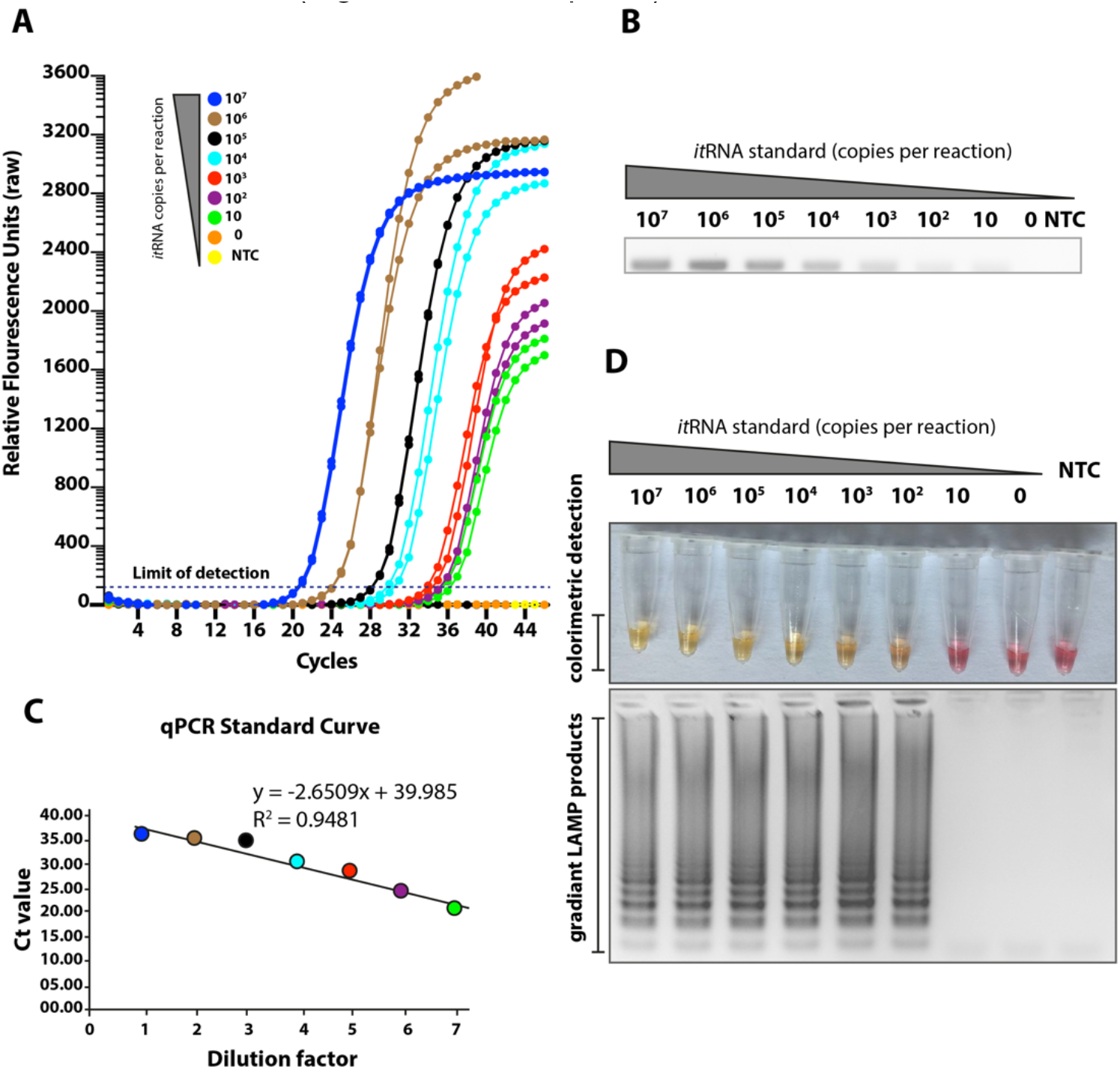
Sensitivities of the LAMP assay. (**A**) Seven different dilutions of in vitro transcribed RNA were run for quantitative measurement using qRT-PCR. Relative fluorescence units show a gradient decrease in signals. (**B**) The corresponding PCR products on the electrophoresis gel (**C**) The qRT-PCR standard curve based on the Ct value and dilution factor. (**D**) The serially diluted synthetic RNAs were run in the LAMP assay and colour change represents positive (yellow) or negative (pink). The lower panel shows the LAMP gradient products.

### 3.3. Cross-reactivity of the novel LAMP assay when tested against other respiratory and medically important viruses

The SARS-CoV-2 embraces genetic and phenotypic features of several common cold coronaviruses and other viruses of the respiratory tract. Owing to high genetic similarity (up to 96% at nucleotide levels) and common respiratory specimen for clinical identification of CoVID-19 patients, we aimed to investigate any non-specific amplification in the LAMP assay. In order to demonstrate the specificity of the LAMP assay, we used pathogens belonging to 5 families of the most important medical and respiratory viruses. As shown in the Figure 3A, the qRT-PCR specifically detected only the SARS-CoV-2 and this was confirmed by Gel-red staining of amplified products (Figure 3B). Consistently, no cross-reactivities were observed with the LAMP in both colorimetric detection (Figure 3C, upper panel) or electrophoreses (Figure 3C, lower panel). Collectively, a highly specific detection of SARS-CoV-2 was observed for primers set using either of the assays.

**Figure 3:**
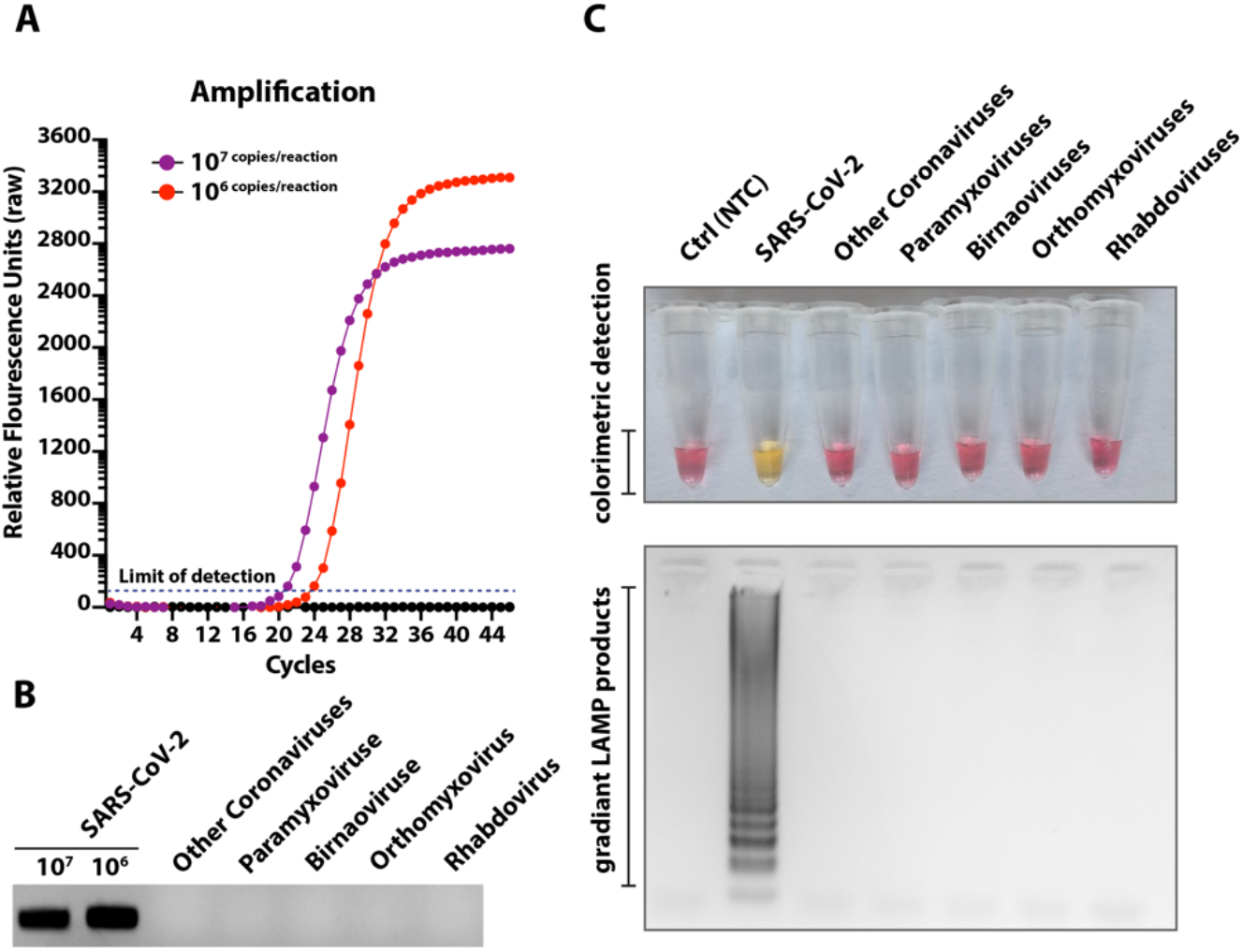
Specificity of the LAMP assay. (**A**) RNA extracted from different medically or respiratory important viruses as well as two dilutions of synthetic RNA were run for qPCR. (**B**) Corresponding PCR products were run on gel to demonstrate specificity. (**C**) Similar to qRT-PCR, extracted RNA were run in the LAMP assays. The top panel indicates the colorimetric detection of LAMP positive/negative reactions and the lower panel show the electrophoresis of the corresponding LAMP products.

### 3.4. Temporal investigations of the LAMP assay and its impact on the limit of detection

One of the major advantages of LAMP is its robustness. In order to determine the optimal time required for sufficient amplification of targeted genes, *in vitro* transcribed RNA was used as a template in 30 minutes assays and assessed every 5 minutes post-start of the reaction. The change in colour was monitored visually by the naked eye. As shown in Table 1, under equivalent conditions similar results were obtained from between 20-30 minutes of amplification. Therefore, to improve sensitivity and avoid missing any weak positives, 30 min was selected as the optimal visual interpretation time for the results.

**Table 1.** Reaction times to visually detect LAMP positivity

| Time in minutes (after start of the LAMP reaction) | *In vitro* transcribed RNA dilution (copies/reaction) |  |  |  |  |  |  |  |  |
| --- | --- | --- | --- | --- | --- | --- | --- | --- | --- |
|  | 10<sup>7</sup> | 10<sup>6</sup> | 10<sup>5</sup> | 10<sup>4</sup> | 10<sup>3</sup> | 10<sup>2</sup> | 10 | 0 | NTC |
| 05 | - | - | - | - | - | - | - | - | - |
| 10 | - | - | - | - | - | - | - | - | - |
| 15 | + | + | + | + | - | - | - | - | - |
| 20 | + | + | + | + | + | + | - | - | - |
| 25 | + | + | + | + | + | + | - | - | - |
| 30 | + | + | + | + | + | + | - | - | - |

While the change in colour, reflective of a positive reaction, could be detected as early as 20 minutes post-start of the reaction at lower copy number, subjective variabilities may result in erroneous interpretation, especially in colorimetric based diagnostic assays. To propose an automated imaging, processing and interpretation of the LAMP based results, we developed a user-friendly device and furnished it with an artificial intelligence based automatic interpretation algorithm.

### 3.5. Manufacture of an isothermal nucleic acid amplification device with colorimetric detection features

A device (Figure 4A) was built with many off-the-shelf electronic components and custom flexible resistive heating elements (5W, NEL, UK), and specially designed aluminium heating blocks. Raspberry Pi (RPi) was used to control the device. The one wire interface of the RPi was used to connect ten digital temperature sensors (DS18B20, Maxim Integrated, USA) positioned directly on the PCB boards to monitor heater block temperature changes and provide feedback control. The specially designed aluminium heater blocks to hold 200 µl PCR tubes and the lid heater to prevent condensation were attached directly on top of the surface mount temperature sensors on the respective PCBs with a heat transferring adhesive (TermoGlue, Termopasty Grzegorz Gasowski, Poland). The flexible resistive heating elements were also attached to the heater blocks. To circumvent the need for specialised docks and eliminate user interpretation of the colorimetric results, a Raspberry Pi Camera (RPi Camera) was used. Eight LEDs (LW T733, Osram, Germany) were assembled on the top side of the lid mount PCB to shine light directly into the reaction tubes to achieve consistent lighting within the device. All the above components were assembled into a 3D printed enclosure (14.3 x 10.8 x 6 cm) specially designed with openings to access to the USB and TCP/IP ports of the RPi. A 20,000 mAh power bank (Anker Power Core, Anker, China) with two 5V, 2A output was used to power the device. A Python based control software was used to control the heating, image the progression of the LAMP assay and store the ‘time-lapse’ images and temperature data within a specified folder. The user can initiate a test by either connecting to a screen via the HDMI port or through simply pairing the device with the mobile app via Bluetooth and selecting the required diagnostic assay.

**Figure 4:**
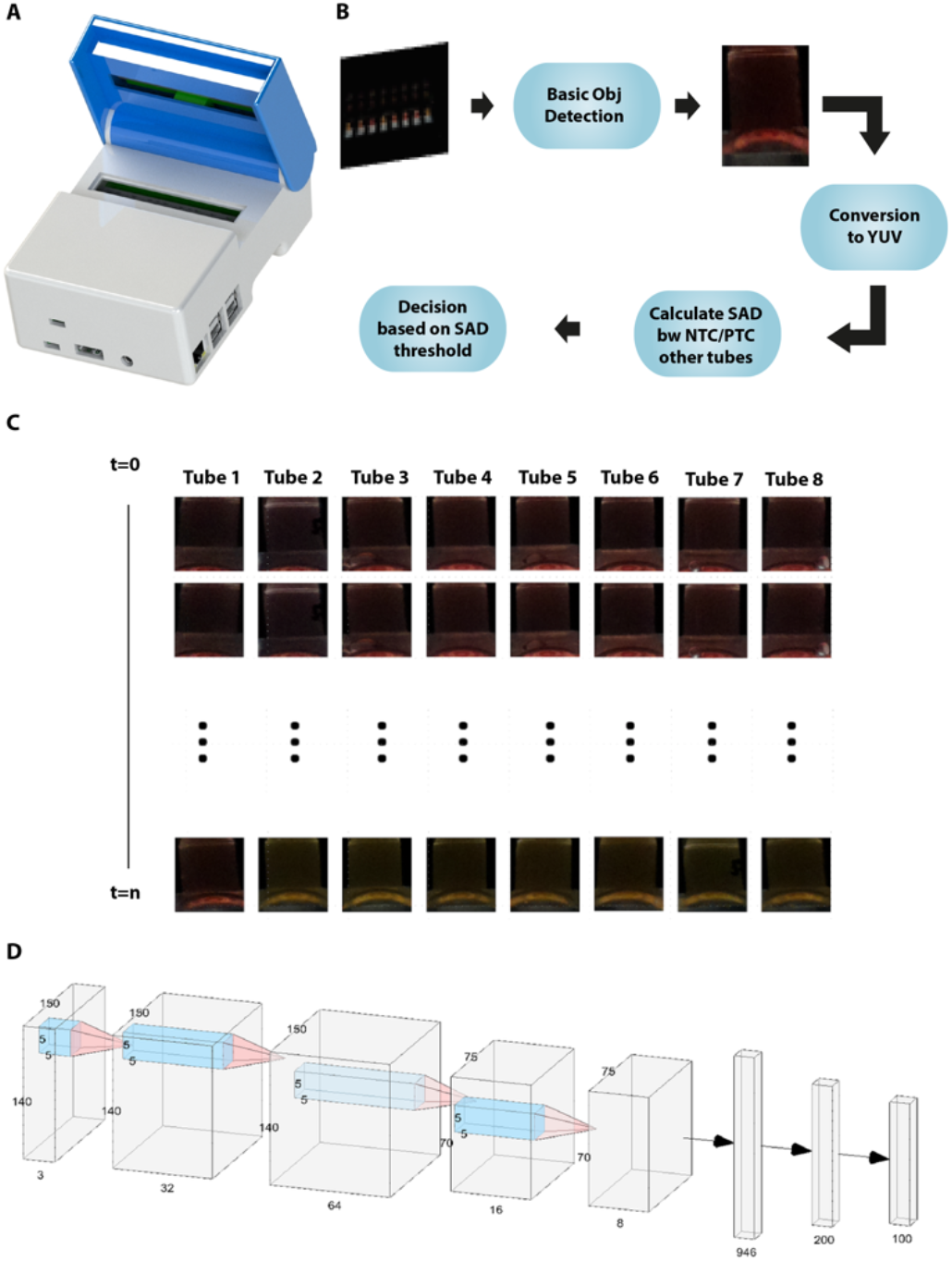
Fabrication and processing of LAMP data for enhanced detection of SARS-CoV-2. (**A**) Exterior of a smart diagnostic device (**B**) Description of the AI-assisted algorithm and image processing. (**C**) Pipeline to process images and extraction of colorimetric information. (**D**) Schematic outlining the training of the network for image processing.

### 3.6. Automated image acquisition and processing through a template matching-based algorithm

The LAMP assay (8 separate tubes) was remotely started to initiate heating to 65°C. Images of those test tubes were captured using the inbuilt RPi Camera for every 20 seconds and were saved in the RPi in the RGB format. Each individual image (90 images in total) consisted of 8 frames for each tube with a black background and these were analysed for 30 minutes. As the tube area exposing colour changes was fractionally small compared to its background, we first extracted each targeted tube frame from the image before applying an image processing algorithm. In order to process these extracted frames, a reference tube was selected as a template, and a template matching algorithm [25] was applied to extract all tubes from the first image. The rationale for the template matching was to search and find the location of a template image in a larger image. It simply slides the template image over the input image to perform the 2-dimensional convolution and compared values to get the maximum overlap to decide the exact similar areas. Assuming that positions of the test-tube do not change over the time of an experiment, images were cropped in an experiment to obtain the tube frames from the entire image. These crops are then saved into a 2-dimensional array for RGB colour space (see equation below). Once extracted, RGB format images were converted to YUV format using the following transformation [26] to avoid diffraction and lighting variabilities in different images.

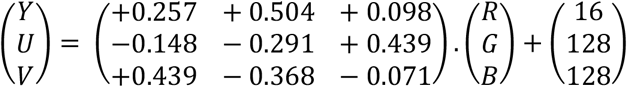

In YUV colour space, the Y channel represented the luminance of the colour, while the U and V channels represented the chrominance (Figure 4B). Separating the luminance from the chrominance reduced the effect of light changing and shadow noises in each tested tube [27]. Finally, the chrominance (U, V) channels from the YUV image were considered for image processing. The chrominance (U, V) values of those extracted test tubes were compared with reference orange test tubes in positive control and reference pink image in negative control test tubes to calculate the sum of absolute difference (SAD) for each of the pixel values. After experiments with two images set and fine tuning the threshold values manually, a SAD threshold value was achieved which provides 100% accuracy in the separation of COVID-19 positive and negative samples.

### 3.7. Artificial intelligence-assisted rapid detection of colour changes associated with the LAMP reaction

Deep learning is a subdomain of AI which doesn’t require any domain knowledge to work. However, it learns hidden patterns from examples present in the dataset. A deep learning Convolutional Neural Network (CNN) [28] architecture was proposed with the bespoke 8 layers deep mode as shown in Figure 4C. It consisted of four convolutional layers followed by 2 dense and an output layer. Binary cross-entropy was used as a loss function for optimizing this CNN model.

For the training of the network, the dataset with 360 images was shuffled and then split into 9:1 proportion (Figure 4D). 90% of the data was used to train the network and the remaining 10% was exploited to check how the network behaved on seeing a new image. Training a dataset requires loading numerous images into the memory in a single operation which is an expensive process. Therefore, a data generator was implemented that read the data in batches from the dataset directory and fed it to the model. After multiple experiments, it was observed that the network converged after 6 cycles (epochs) through the dataset. Therefore, we ran an experiment for only 6 epochs to decrease the probability of overfitting. In addition, an additional set of 108 test-tube crops was used to validate the network. The best performing network resulted in an accuracy of 98% in classifying tubes based on their colours (images with better light).

In order to assess the temporal impact of the AI-assisted detection of colour changes (indicative of amplification), the RT-LAMP reaction was run with 3 previously confirmed positive and negative patient samples in addition to known positive and negative controls. Colour changes were assessed every 5 minutes until the completion of the LAMP reaction at 30 minutes. Gradual colour changes were detectable with the naked eye as early as 20 minutes post-start of the reaction (Figure 5A). Corresponding samples were run on the newly developed device and temporal and real-time colour changes were monitored as described earlier. Depending upon the viral load in the test sample, a clear colour change was detected and calibrated as early as 20 minutes using device operated processing of the data (Figure 5B). Once the positive test control was identified as positive, the test was be stopped, and the results were returned, thus reducing the waiting time and power consumption.

**Figure 5:**
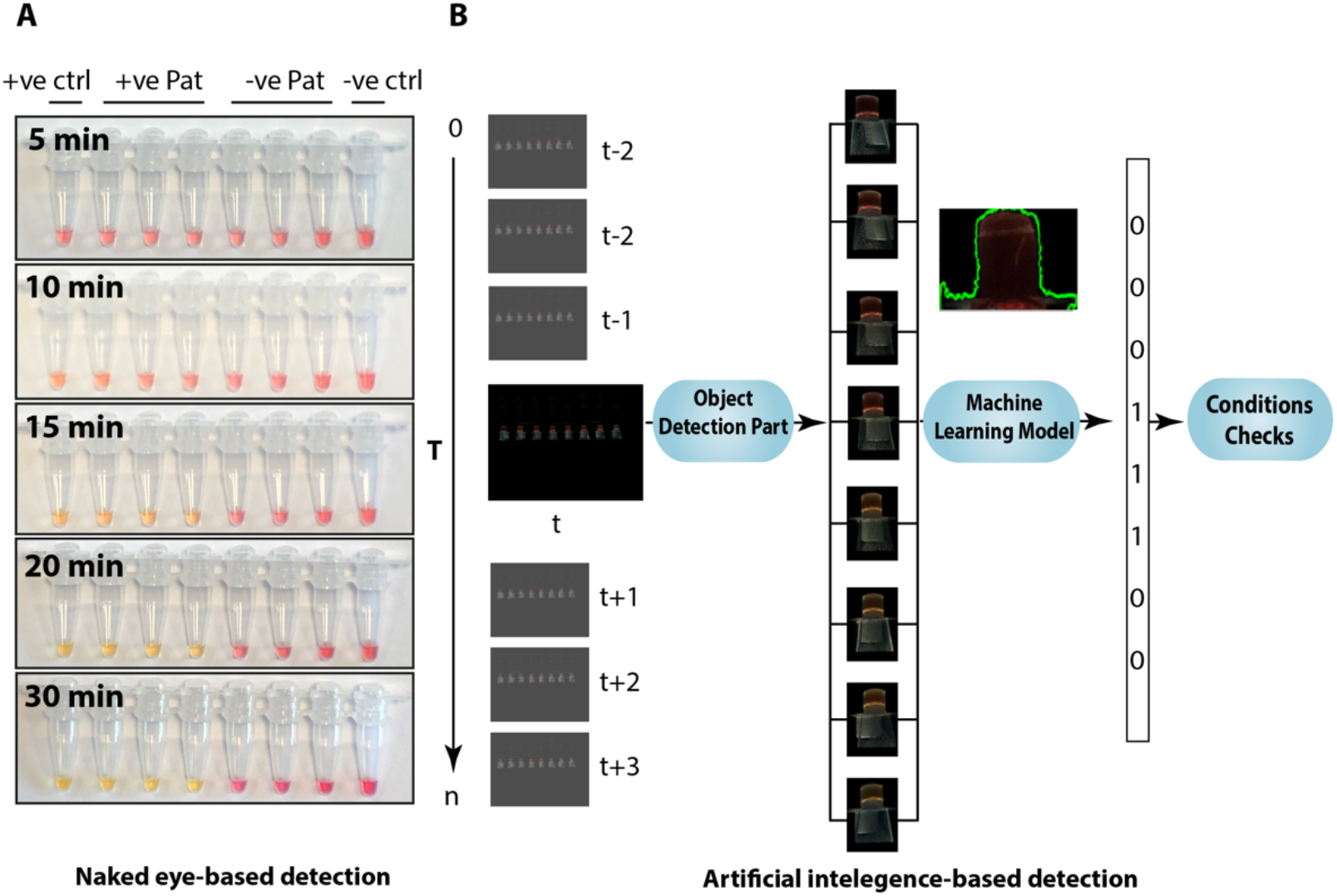
Conventional and AI-assisted interpretation of LAMP results. (**A**) Temporal analysis of known positive and negative patient samples from 360 images taken from RPi for visual interpretation of LAMP results. (**B**) Interpretation of corresponding patient samples by the AI-assisted LAMP results.

As shown in Fig 5B, a template matching algorithm was applied to extract test tubes from images the CNN model was applied to the extracted tubes to train the model. The CNN model was used as a machine learning algorithm to identify colours for each image taken throughout the experiment. Images taken at time tare marked as ‘t’ in the Fig 5B. Once the positive control test and the negative control test gave the correct results consecutively on three occasions, the LAMP based test was be stopped, and the results returned. This approach reduces waiting times for the results and power consumption.

### 3.8. Validation of ai-LAMP and comparative performance in clinical settings

In order to assess the field application of the optimized assay, we applied the ai-LAMP to purified RNA spiked with miR-cel-miR-39-3p from CoVID-19 patients. A total of 199 swab samples were collected from CoVID-19 clinically suspected patients during routine COVID-19 screening at the Royal Lancaster Infirmary (RLI), University Hospitals of Morecambe Bay NHS Foundation Trust UK. The extracted RNA from swab samples were run in parallel for ai-LAMP and two WHO/PHE recommended qRT-PCR targeting the RdRP and N genes of the SARS-CoV-2. This parallel assessment facilitated the assessment of the comparative performance of the ai-LAMP.

The RdRP gene-based qRT-PCR detected a total of 67 positives and 132 negatives in a cohort of 199 patients (Figure 6A). In contrast, a higher number of positive (n=88) and lower numbers of negative (n=111) were detected by the qPCR which targeted the N gene (Figure 6B). Interestingly, the ai-LAMP detected a total of 126 positive samples which constituted several times higher than RdRP and N gene-based qPCR, respectively. Comparative analysis of these three molecular detection assays revealed 58 total true positives (TP), 09 false negatives (FN), 64 true negatives (TN), and 68 false positives (FP) in RdRP-based qRT-PCR compared to RdRP-based LAMP (Figure 6A). Similarly, upon comparative analysis of the N gene-based qPCR and RdRP-based LAMP, we observed a total of 74 TP, 14 FN, 59 TN, and 52 FP (Figure 6B).

**Figure 6.**
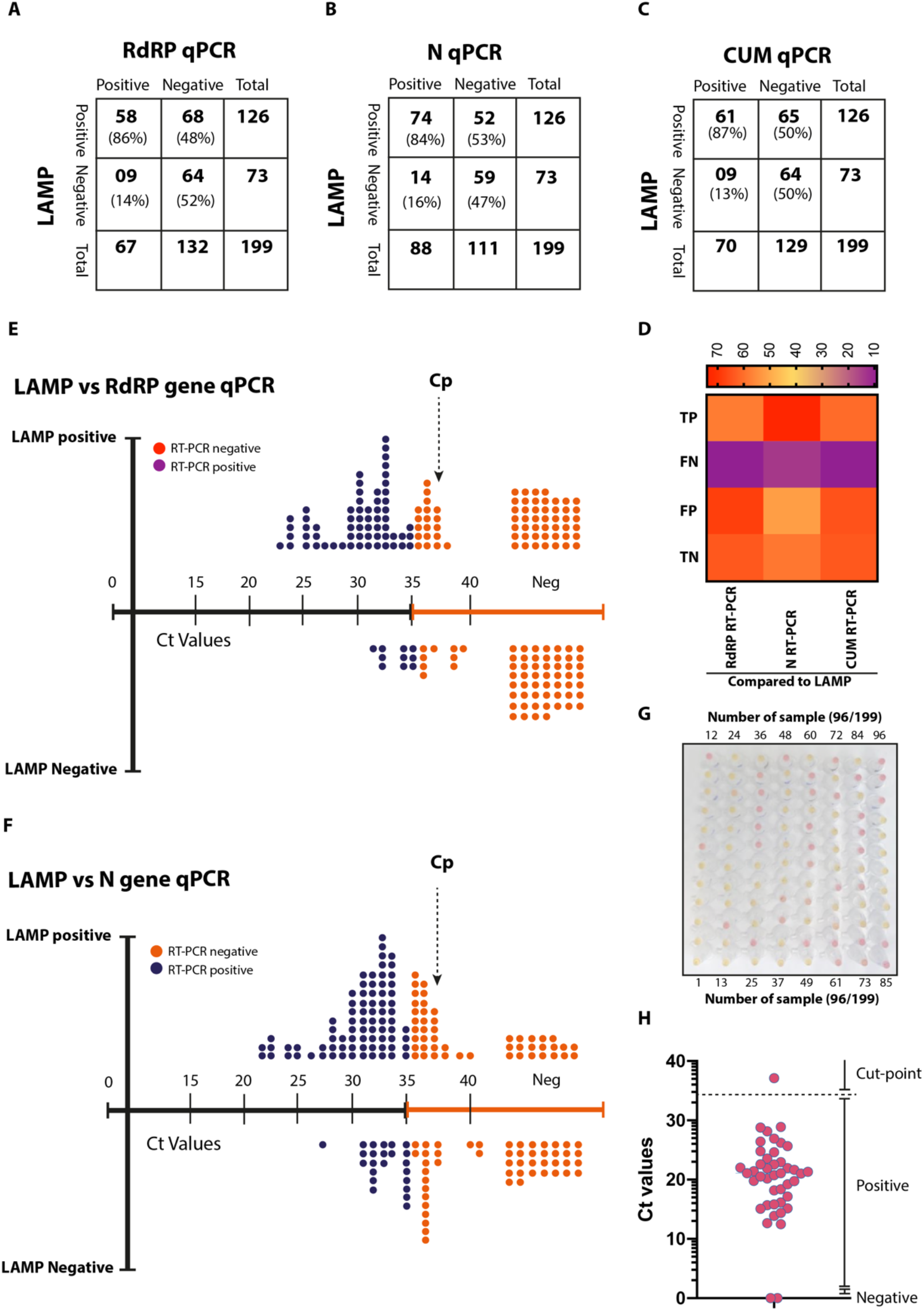
Clinical validation of ai-LAMP. (A**-C**) Comparative sample positivity between LAMP and RdRP qRT-PCR (**A**), LAMP and N qRT-PCR (**B**), LAMP and CUM qRT-PCR results (**C**). (**D**) The heatmap indicate the relative positive and negative samples among three assays. (**D**) Linearity chart comparing the LAMP positive/negative samples and their detection based on the RdRP gene-based qRT-PCR. (**F**) Linearity chart comparing the LAMP positive/negative samples and their detection based on the N gene-based qRT-PCR. (**G**) Naked eye detection of the first 96 samples out of the total 199 patients’ samples were processed. (**H**) Recovery Ct values of the miRNA from spiked before RNA extraction.

In the current clinical settings, a qRT-PCR targeting two genes (N and RdRP) was conducted to conclusively identify CoVID-19 positive cases and this assay is referred as cumulative (CUM) qRT-PCR. In this scenario, a sample would be considered as positive only if a Ct value of =/> 35 was detected in both N and RdRP-gene based qRT-PCR. Using this approach, we noticed a total of a 70 positive and 129 negative samples and an improved true positive (n=61), false negatives (n=09), true negatives (n=64), and false positives (n=65) limits (Figure 6C). Taken together, the cumulative comparative picture of the qPCR and ai-LAMP has identified a superior detection of positive cases (Figure 6D). In order to confirm this detection performance, all ai-LAMP amplification products were visualised by electrophoresis (data not shown), further confirming the aided-detection and improved implication of ai-LAMP in field conditions.

Next we determined the detection limit of the ai-LAMP in direct correlation with the standard Ct values of the qPCRs. Plotting of ai-LAMP positive and negative data against the linearity of the Ct values revealed that ai-LAMP carried Cp (cycle number at detection threshold) of up to 37 Ct determined by the RdRP gene-based qPCR (Figure 6E and Supplementary Table 1) or N gene qRT-PCR (Figure 6F and Supplementary Table 1) of the SARS-CoV-2. This detection was approximately 2 Ct values higher than the detection limit of the standard qPCR. Analysis of the first 96 samples, run in parallel for the ai-LAMP, (Figure 6G and Supplementary Table 1) showed a clear demarcation of the positive and negative samples in the RdRP-gene based ai-LAMP. In order to rule out the quantitative recovery from spiked miRNA, a qRT-PCR was run on 40 randomly selected RNA samples [23]. Based on the quantitative Ct values for miRNA, all samples showed a marked recovery except a single sample where a low detection of the miRNA was identified (Figure 6H, Supplementary Table 1 and Supplementary Figure 1).

Collectively, these data highlight the improved specificity and sensitivity of the AI-assisted LAMP assay compared to the naked-eye interpretation of the LAMP result, thus enhancing the timely and automated detection, and interpretation of the assay results (see supplementary data).

## 4. Discussion

The SARS-CoV-2 is now a global pandemic, over 216 countries are currently reporting active infections and the number of daily infections and deaths is continuing to rise, especially in the Americas and South East Asia through a series of multiphasic spread [29]. Currently, there is no licensed vaccine or registered drugs, leaving timely identification of CoVID-19 patients, contact tracing and isolation of positive contacts as the most effective means of containing the pandemic. Among different molecular diagnostic chemistries, LAMP technology provides a promising approach for the rapid and reliable detection in resource-limited settings [17]. Recently, the LAMP technology has been widely applied for the identification of West Nile virus, influenza virus, yellow fever virus, Marburg virus, Ebola virus, Zika virus, and other myriads of viruses [30-35]. Here, we demonstrated the specificity, sensitivity and utility of a novel ai-LAMP assay for SARS-CoV-2.

The genome of SARS-CoV-2 is approximately 30kb in size with a coding capacity of 9860 amino acids. All of the β-coronaviruses encodes for structural (replicases, S, E, M and N) genes in the order of 5’ to 3’ in the positive sense genome [5, 36, 37]. A range of qRT-PCRs have been proposed and are referred by the World Health Organization [29; https://www.who.int/emergencies/diseases/novel-coronavirus-2019/technical-guidance/laboratory-guidance] for diagnosis of SARS-CoV-2. While diagnostic assays can be designed on the most conserved region of the viral genome, most of the routinely applied RT-PCR and RT-LAMP have been targeting the S, N, RdRP, E and ORF1a/b genes mainly due to their high level of transcription and abundance in expression compared to other genes of the SARS-CoV-2 [5, 6, 38]. For the detection of SARS-CoV-2, Chan et al., [23] have targeted and developed a standard RT-LAMP with LoD of 11.2 RNA copies/reaction using *invitro* RNA transcripts. Yan et al., [16] have adapted the ORF1ab to developed RT-LAMP assay with a detection limit of sensitivities of 2×10^1^ copies per reaction. The majority of these diagnostic assays carry a high level of sensitivity, specificity and repeatability; however, these primarily lack the clinical validation and/or optimization on the synthetic targets.

In this study, we have developed and evaluated a novel RT-LAMP in one of the most conserved genes (i.e. RdRP) within the SARS-CoV-2 genome. The RT-LAMP was then directly compared with the currently applied routine diagnostic assays to assess the comparative performance. The RT-LAMP assay developed in this study, could detect as low as 100 copies with an *in vitro* RNA transcript. Importantly, the RT-LAMP has detected the SARS-CoV-2 RNA in 68/199 (34%) and 52/199 (26%) additional specimens that were tested negative by the RdRP-based qRT-PCR and N-based qRT-PCR, respectively. These findings are interesting, both clinically and epidemiologically due to the high proportion of asymptomatic and mildly symptomatic cases of CoVID-19. These apparently healthy people have been suggested to be a major sources of virus propagation and the basis of epidemics within the community [39-41]. Therefore, highly sensitive and specific test is needed to identify cases with low viral load. The RT-LAMP detected more positive samples which were otherwise negative by routinely applied qRT-PCR assay. In order to assess the potential false positive identification, we run electrophoreses and sequencing of the N gene. The use of a spiked RNA standard that is not expressed in humans (cel-miR-39-3p) helped to confirm the effective effectiveness of the RNA extraction approach using commercial kits (Qiagen). In addition, we used a fixed total RNA concentration in all experiments allowing for better comparisons across groups.

The main challenges of using the colorimetric approach are background which changes the colour perspective, issues in identifying small changes, bubbles in the test tubes, relatively small area corresponding to colour change and pixel variation due to camera flash and background reflections. The CNN based model has used high-performance computer images to train using these issues and having learned the patterns is able to classify colour, despite the presence of noise. The trained model has successfully moved to Rpi to identify colour changes in test tubes. The study produced 98% accuracy for images taken with better light (Open) and the duration of testing could be dynamically controlled to reduce the length of operating time and heating with a resulting reduction in energy consumption by the device. Despite the SAD based approach resulting in 100% accuracy for the images after 30 minutes, this approach failed with other datasets containing bubbles and different background lights as a different threshold value was produced for each image set. Therefore, a convolutional neural network (CNN) approach was utilised in our experiments to generalize the classification for orange and pink test tubes with different background light and bubbles.

Collectively, our data showed that the newly established ai-LAMP was highly specific for the detection of SARS-CoV-2 RNA from extracted respiratory tract clinical specimens. The application of this novel LAMP assay may be particularly useful for detecting COVID-19 cases with low viral loads and when testing upper respiratory tract specimens (nasal or oral swabs) from patients. Development of ai-LAMP into a multiplex assay which can simultaneously detect other human-pathogenic coronaviruses and respiratory pathogens may further increase its clinical utility in the future.

## Data Availability

All data required has been made available within the manuscript.

## Supplementary Materials

The following are available online at www.mdpi.com/xxx/s1, Figure S1: Optimization and raw data on the LAMP optimization, Table S1: Data on the comparison of the qRT-PCR and LAMP.

## Author Contributions

Conceptualization, M.M., A.F. and W.B; methodology, M.A.R, W.B. and M.M.; software, A.F.; validation, M.T., N.S.C. and M.A.R.; formal analysis, M.A.R, A.P. M.B; investigation, M.M., M.A.R; resources, M.M., A.F. and W.B; writing—original draft preparation, M.M; writing—review and editing, M.A.R, E.C., I.S., J.V., M.E.K, M.Q.A, M.B., A.P., M.B., M.T., N.S.C., R.S., A.B., P.B., W.H., J.B., J.B., H.W., C.W., M.B., R.L.R., W.B., A.F., and M.M; supervision, R.L.R., A.F., W.B., and M.M.; project administration, M.M; funding acquisition, R.L.R., A.F., W.B., and M.M. All authors have read and agreed to the published version of the manuscript.

## Funding

The authors wish to express our sincere appreciation to the BBSRC for allowing us to repurpose the LAMP prototypes produced in the grant BB/R012695/1 to be used for SARS-CoV-2 laboratory testing at The University of Lancaster. We would like to thank the support of BBSRC (BB/M008681/1 and BBS/E/I/00001852) and British Council (172710323 and 332228521) at Division of Biomedical and Life Sciences, Lancaster University, UK. We would also like to thank Brunel University London and the University of Surrey for providing some financial support to rapidly produce these devices.

## Acknowledgments

The authors would like to thank the Electronic Technicians William Schkzamian, Gopalakirishnan Jeysundra and Michael Lateo of Brunel University London for their efforts to travel to the University with special permission during the early lockdown period to produce eight laboratory prototypes within 5 days. We thank the Microbiology Department, University Hospitals of Morecambe Bay for access to anonymised patient samples and acknowledge the support of BLS Lancaster University Technicians throughout the lockdown period. We would like to thank Dr Derek Gatherer, Lancaster University, in aligning large SARS-COV-2 genome sequences.

## Conflicts of Interest

The authors declare no conflict of interest. The funders had no role in the design of the study; in the collection, analyses, or interpretation of data; in the writing of the manuscript, or in the decision to publish the results.

## Notes

### Competing Interest Statement

The authors have declared no competing interest.

### Author Declarations

This study was conducted in accordance with the University Human ethics guidelines and received a favourable review from the Faculty of Health and Medicine Research Ethics Committee (FHMREC) of Lancaster University - reference number FHMREC19112.

